# Estimation of fatality rate in Africa through the behavior of COVID-19 in Italy relevance to age profiles

**DOI:** 10.1101/2020.06.21.20129049

**Authors:** Lambert Nzungize, Diane Umuhoza, Yongdong Dai, Stech A E. Nzaou, Mohammed Asaad, M. A. Abokadoum, Ulrich Aymard Ekomi Moure, Jianping Xie

## Abstract

The emergence and pandemic of COVID-19 has rapidly become a global concern. In Italy, on 27 March 2020, there were 8165 deaths and 80539 confirmed cases of COVID-19. Demographic situations, like age profiles is reported to be the cause of high case fatality rate (CFR) in Italy. In Africa, the COVID-19 pandemic has not yet grasped epic proportion, but the estimation of CFR is still needed. We compared the CFR observed in Italy with the age profiles in 46 Africa countries and 2 territories which are already confirmed COVID-19 case. The estimation of the CFR in Africa ranges between (1.0%-5.4%) while in Italy is 10.1%. The five highest CFR countries and territories in Africa are Reunion (5.4%), Mauritius (5.1%), Tunisia (3.9%), Seychelles (3.8%) and Morocco (3.3%). The last three countries with low CFR are Uganda (1.0%), Zambia (1.1%) and Angola (1.1%). The observed difference is related to the age profiles.

## Introduction

Africa continent holds 54 countries with 1.3 billion of the population with a median age of 19.7 years old [1]. Up to now around 46 countries and 2 territories already have confirmed 2419 people infected and 39 deaths from COVID-19 according to World Health Organization (WHO) situation report [2]. Each country in Africa declare its own decisive public health measures according to their local epidemiological situation amid COVID-19 such as bans on public gathering, canceling conference or sports events, closing borders, bans travel restrictions, shutting downs schools, self-quarantine to enhance social distancing as WHO and health experts recommended [3] to slow the country’s rate of COVID-19 transmission. The widespread of COIVD-19 has infected over 509164 people worldwide by 27 March 2020 [2]. People with pre-existing conditions like cardiovascular disease, diabetes, chronic respiratory disease, hypertension and cancer are more susceptible to COVID-19 [4]. We hypothesized that if the age structure is a crucial key of the high case fatality rate in Italy from COVID-19, what can be happened in Africa based on the demographic profiles. Italy is one of the Mediterranean nations of 60 million and the second country with 23% of the population over 65 years old after Japan [5]. Italy has reported high CFR of 10.1% (8165 deaths/8053 cases) [2], while 37.8% of infected people are between 51-70 years old and 35.8% of infected people are above 70 years old [6]. Therefore, the median age of people infected by COVID-19 is 62 years old where 57.2 % are male and 42.8% female [6].

## Results and Discussion

We chose 46 countries and 2 territories with at least one COVID-19 case confirmed in Africa by 27 March 2020 [2]. The significative difference of the young age population in Africa relatively describes the reason for low fatality rate (Fig.1) versus Italy where COVID-19 is more fatal in older patients [7]. During the epidemic, it’s essential to estimate the case fatality ratio [8]. The estimation of CFR based on age profiles using data in Italy is not confidently effective to control COVID-19 as shown in Fig 2, however the near future estimation is necessary based on actual situation of CFR in Italy (Fig. 3) as reported 10.1% [2]. Here we propose that Africa countries should evaluate and enforce public health measures to facilitate public health interventions and dwindle the CFR (Fig. 3). We found that, in Italy the age profiles is the first reason which is leading to the high CFR from COVID-19, furthermore, now Italy is classified as hyperendemic country, the top country with antibiotics resistance deaths in Europe [9], which can be the second reason to cause the pneumonia infection like COVID-19 to be more lethal. In turn, Africa is the niche of two epidemics such as HIV (human immunodeficiency viruses) which weakens a person’s immune system and tuberculosis (TB) as a lung disease associated with AMR (antimicrobial resistance) [10-12]. Those dual epidemics might worsen the situation of COVID-19 in Africa. Hence, strict public health measures are required to prevent the spread of COVID-19 to the people infected with HIV/TB. Africa countries with weak health care system or deficiency of testing resources to the imported cases may arise an overestimation of the CFR to the population in close territory or region (western, southern, northern, or central) as shown in Table 1.

**Figure 1.**
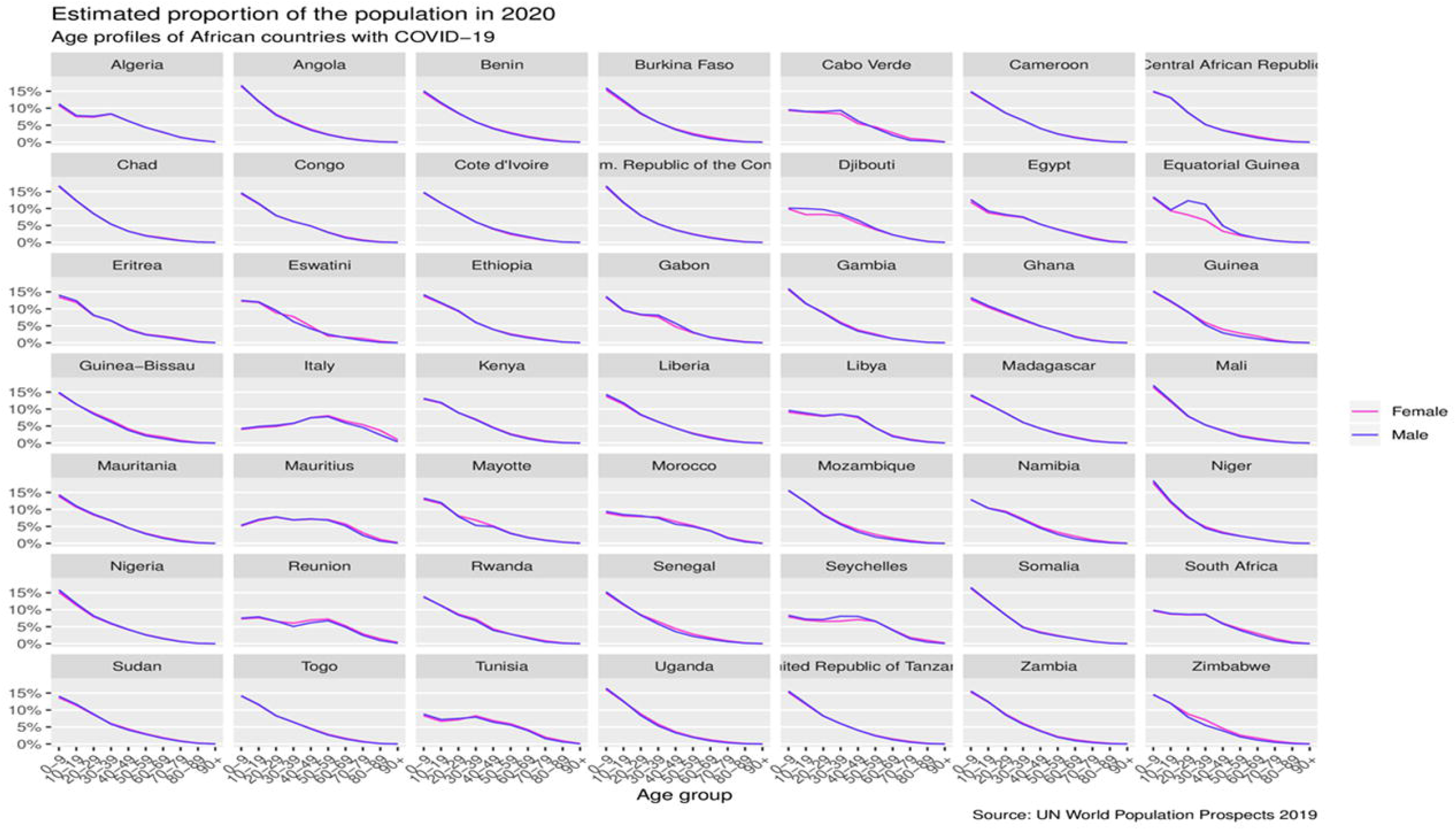
Trends in levels of the population with an estimated proportion of the population in 2020 by age group and the line show the rates of sex (female and male) by each specific country.

**Figure 2.**
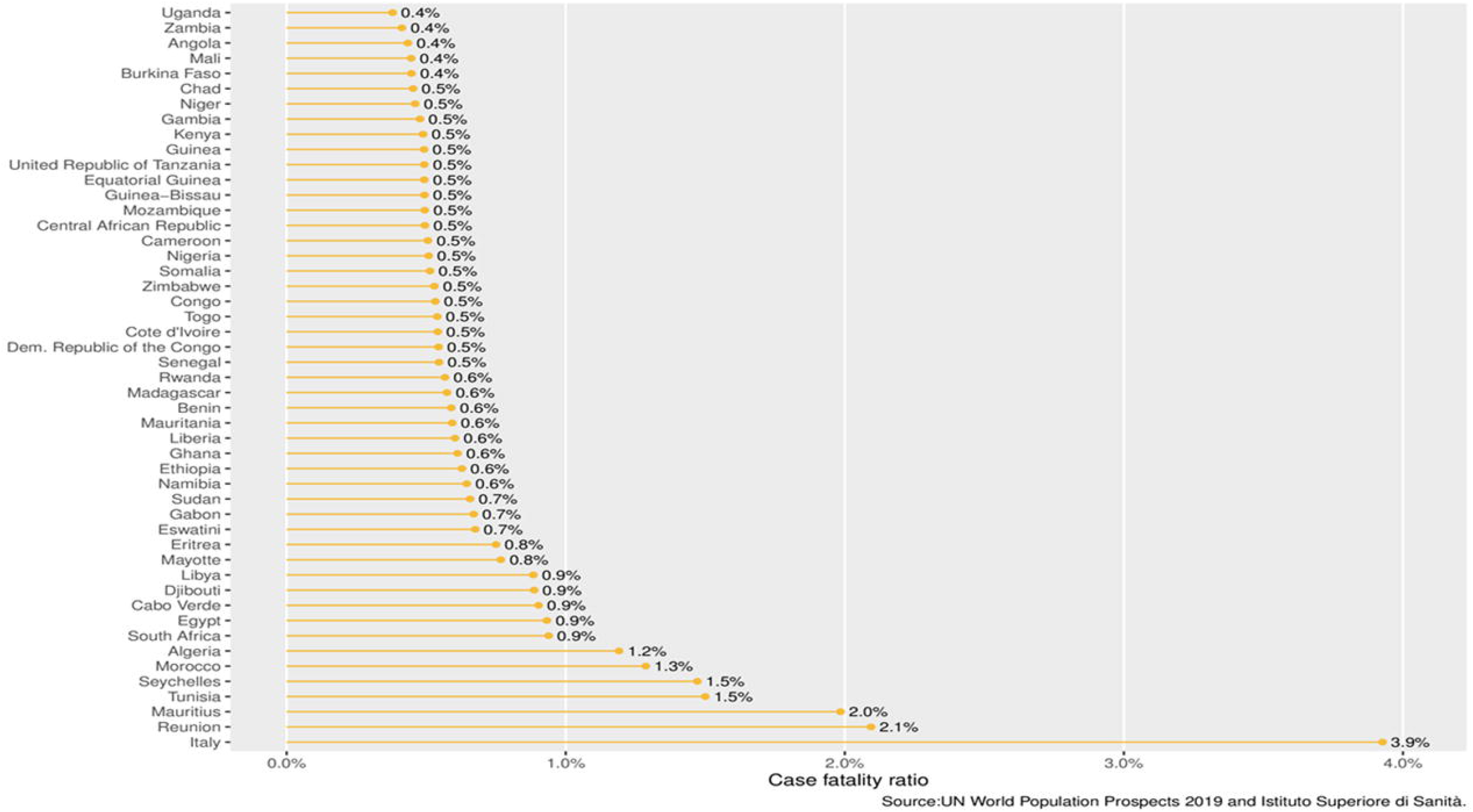
Different age structures among countries can alter significantly the overall case fatality ratio based on Italian age profiles.

**Figure 3.**
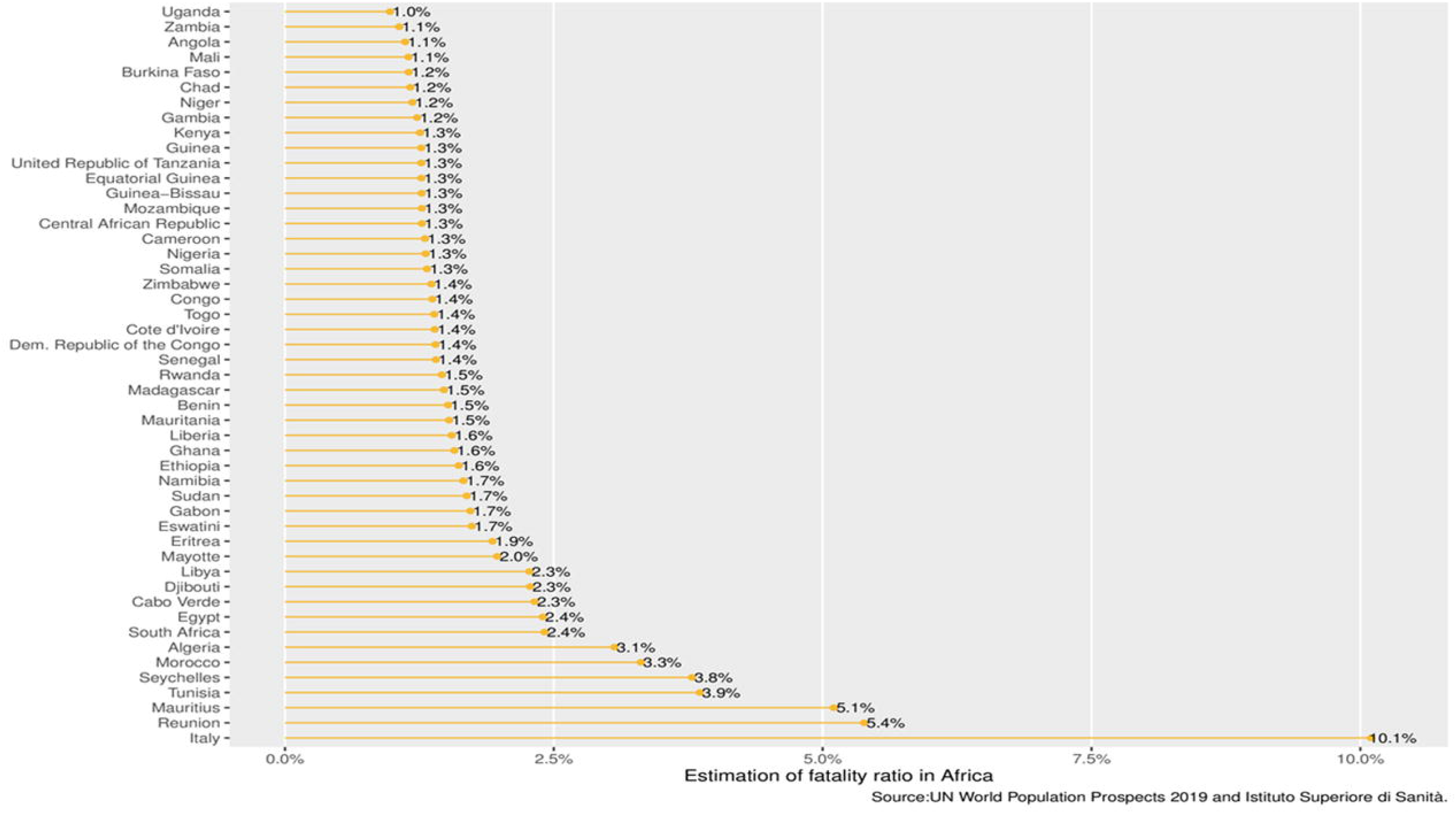
Estimation scaled of CFR of 10.1 % observed in Italy (27 March 2020) implemented to Africa countries with confirmed COVID-19 cases by 27 March 2020.

**Table 1.** Africa countries with COVID-19 per region

| **Region** | **Country** |
| --- | --- |
| Central | Cameroon, Central Africa Republic, Chad, Congo, Dem. Republic of Congo, Equatorial Guinea, Gabon. |
| Eastern | Djibouti, Eritrea, Ethiopia, Kenya, Madagascar, Mauritius, Rwanda, Tanzania, Uganda. |
| Northern | Algeria, Egypt, Libya, Mauritania, Morocco, Tunisia. |
| Southern | Angola, Eswatini, Mozambique, Namibia, South Africa, Zambia, Zimbabwe. |
| Western | Benin, Burkina Faso, Cabo Verde, Côte d'Ivoire, Gambia, Ghana, Guinea, Guinea Bissau, Liberia, Mali, Niger, Nigeria, Senegal, Togo. |
| Territory | Mayotte, Reunion. |

**ource:** Africa CDC 2020 [15]

## Conclusion

It is reasonable to wonder why the transmission of COVID-19 is slow in Africa, we found that the age distribution in Africa by each country explain the low fatality rate when compared to the age profiles in Italy. Thus, African leaders and the society should practice strict public health measures and special attention should be oriented to elder people in Africa countries/territories including Reunion (5.4%), Mauritius (5.1%), Tunisia (3.9%), Seychelles (3.8%) and Morocco (3.3%).

## Data Availability

No supplementary data

## Data analysis

Data on all COVID-19 cases in Italy were obtained from the Italian National Institute of Health (Istituto Superiore di Sanità, ISS) which hosting the overall information of people infected by COVID-19 via surveillance system throughout the country [6], and African region data were obtained from WHO coronavirus situation report [2], reported on 27 March 2020. We used R language to estimates the fatality ratio of COVID-19 between Africa countries through various R packages such as wpp2019 of world population data [13], tidyverse, scales and we use similar project code with little modification [14].

## Acknowledgments

Special thanks go to the team members who maintain the update of database of COVID-19 (nCov2019 and coronavirus).

## Authors contribution

All authors have seen and approved the final manuscript.

## References

1. Worldometers. Population of Africa 2019. Available at: https://www.worldometers.info/world-population/africa-population/. Accessed 24 March.

2. WHO. Coronavirus disease 2019 (COVID-19) Situation Report-67. Available at: https://www.who.int/docs/default-source/coronaviruse/situation-reports/20200327-sitrep-67-covid-19.pdf?sfvrsn=b65f68eb_4. Accessed 27 March.

3. WHO. Global surveillance for COVID-19 caused by human infection with COVID-19 virus. Available at: https://apps.who.int/iris/bitstream/handle/10665/331506/WHO-2019-nCoV-SurveillanceGuidance-2020.6-eng.pdf. Accessed 24 March.

4. WHO. COVID-19 Coronavirus disease (COVID-19) advice for the public: Myth busters. Available at: https://www.who.int/emergencies/diseases/novel-coronavirus-2019/advice-for-public/myth-busters. Accessed 24 March.

5. Haider F. Countries With The Largest Aging Population In The World. Available at: worldatlas.com/articles/countries-with-the-largest-aging-population-in-the-world.html. Accessed 24 March.

6. Sanità ISd. Integrated surveillance of COVID-19 in Italy. Available at: https://www.epicentro.iss.it/coronavirus/bollettino/Infografica_27marzo%20ENG.pdf. Accessed 27 March.

7. Onder G, Rezza G, Brusaferro S. Case-Fatality Rate and Characteristics of Patients Dying in Relation to COVID-19 in Italy. Jama 2020.

8. Ghani AC, Donnelly CA, Cox DR. Methods for estimating the case fatality ratio for a novel, emerging infectious disease. American journal of epidemiology 2005; 162(5): 479–86.

9. La Fauci V, Alessi V. Antibiotic resistance: where are we going? Annali di igiene: medicina preventiva e di comunita 2018; 30(4 Supple 1): 52–7.

10. Telisinghe L, Charalambous S, Topp SM. HIV and tuberculosis in prisons in sub-Saharan Africa. Lancet (London, England) 2016; 388(10050): 1215–27.

11. Bernabé KJ, Langendorf C, Ford N, Ronat JB, Murphy RA. Antimicrobial resistance in West Africa: a systematic review and meta-analysis. International journal of antimicrobial agents 2017; 50(5): 629–39.

12. Wangai FK, Masika MM, Lule GN. Bridging antimicrobial resistance knowledge gaps: The East African perspective on a global problem. PLoS One 2019; 14(2): e0212131.

13. UN. World Population Prospects: The 2019 Revision. Available at: http://population.un.org/wpp. Accessed 24 March.

14. statistics Fr. Impact of a country’s age breakdown on COVID-19 case fatality rate. Available at: http://freerangestats.info/blog/2020/03/21/covid19-cfr-demographics. Accessed 24 March.

15. Novel Coronavirus (2019-nCoV) Global Epidemic. Available at: https://africacdc.org/disease-outbreak/novel-coronavirus-2019-ncov-global-epidemic-25-february-2020/. Accessed 27 March

